# Hyperspectral imaging in the emergency department to characterize lower leg edema

**DOI:** 10.64898/2026.09.15.26363173

**Authors:** Karthik Kasi, Cameron May, Hailey Gallegos, Leo Kobayashi, Bevil R. Conway, Joseph R. Pare

## Abstract

Interactions between light and biological tissue could reveal disease-related changes that could be advantageous for rapid diagnosis when applied to whole limbs. Here, we tested the extent to which visible/near-infra-red hyperspectral imaging (HSI) deployed in the emergency department could classify cellulitis, non-cellulitis edema, and healthy lower-leg tissue across skin pigmentation types. We collected HSI data of the lower leg from 83 emergency-room patients and analyzed the calibrated normalized spectra (400-1000 nm) using a machine-learning classification algorithm, evaluated by cross-validation and benchmarked against standard spectral indices (oxygenation, hemoglobin, water, near-infrared perfusion). Classification at patient-level and pixel-level resolution performed comparably well using whole-spectrum and indices, for cellulitis versus healthy (AUC: 0.93 vs. 0.96) and non-cellulitis edema versus cellulitis (AUC: 0.92 vs. 0.89). Full-spectrum analysis substantially improved non-cellulitis edema versus healthy classification (patient-level AUC: 0.72 vs. 0.41; pixel-level AUC: 0.78 vs. 0.50), with no significant effect of skin type on accuracy.

## Introduction

Hyperspectral imaging (HSI) captures dozens to hundreds of narrow light bands, producing a reflectance spectrum at every pixel and enabling quantitative tissue characterization beyond conventional RGB imaging (1)(Figure 1A). The visible/near-infrared range used here (VNIR; 400–1000 nm) is increasingly applied in biomedicine (2, 3), with machine learning expanding analysis of its high-dimensional data (4). Tissue reflectance is shaped primarily by hemoglobin, melanin, fat, water, bilirubin, and β-carotene, whose concentrations vary with genetics, environmental factors, and disease (7, 8). Disease-related changes in skin chromophores may therefore provide spectral signatures that HSI and machine learning can use for diagnosis (Figure 1B). Compared with specialized imaging systems, HSI can be less expensive, easier to operate, more portable, faster, and non-invasive, making it attractive for point-of-care and resource-limited settings (5).

**Figure 1.**
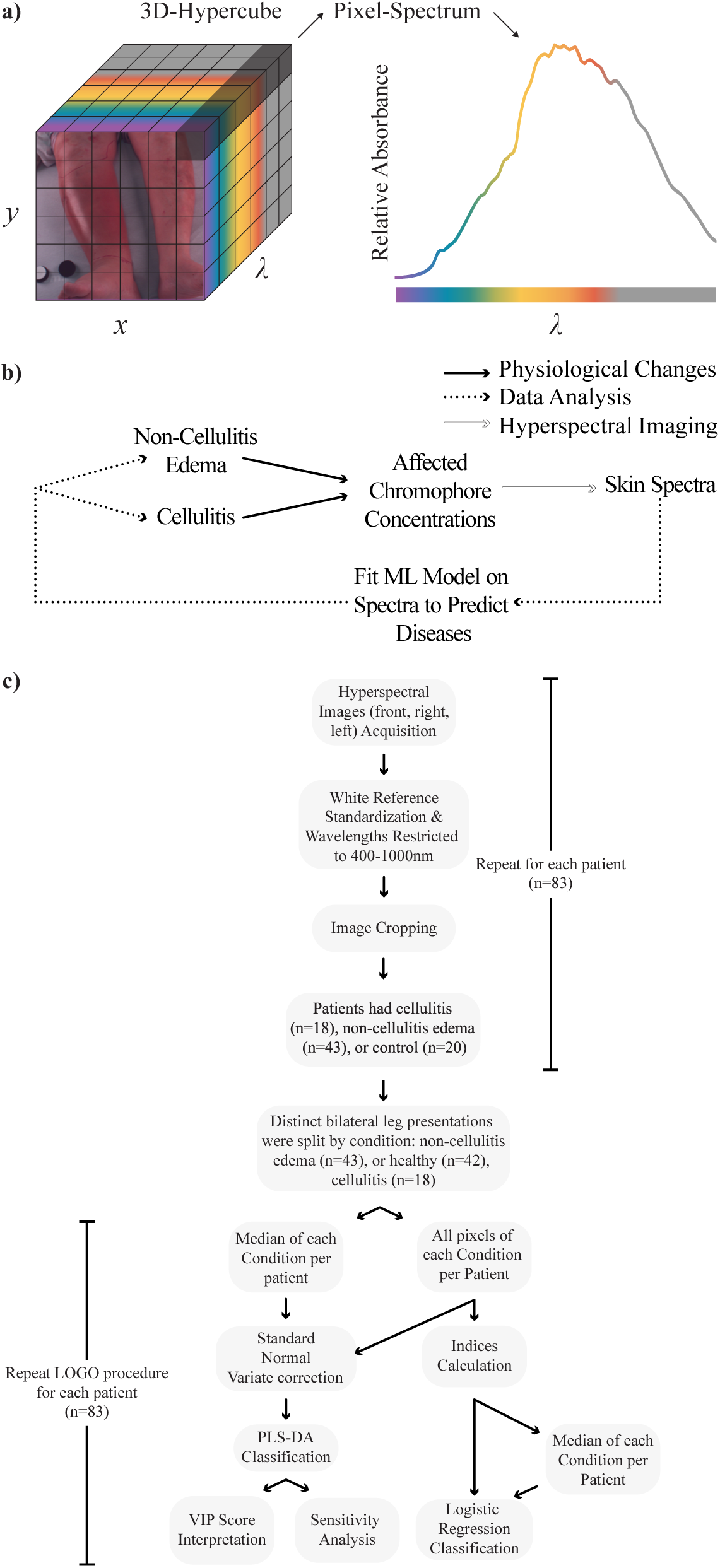
Graphical data analysis overview. A Graphical (left) and plotted (right) representation of HSI data captured from imaging of a study patient’s lower extremities. A hyperspectral image can be conceptualized as 128 different images each at a different wavelength where x and y are spatial dimensions and λ is the wavelength dimension. The visible spectrum is plotted in color, and the infrared region is plotted in grey. B Characteristic changes in skin chromophores allow for prediction of diseases. C Chromophore maps and classification using chromophore indices

As a result, HSI is a natural fit for the emergency department. However, it remains unclear whether spectral measurements are reliable under the variable conditions of this setting. Differences in patient position, imaging angle, distance, and ambient illumination can alter measured reflectance independently of tissue biology, potentially obscuring disease-associated spectral signals (9–11). Skin pigmentation poses an additional challenge because melanin alters baseline reflectance and may obscure erythema or introduce systematic differences in model performance (12). To improve robustness and clinical interpretability, prior clinical HSI studies have reduced the spectrum to indices of tissue oxygenation (StO_₂_), hemoglobin (THI), water (TWI), and near-infrared perfusion (NIR) (13–17). These targeted measures may limit sensitivity to broad baseline differences (such as differences in skin pigmentation), but they also discard information distributed across wavelengths outside their predefined bands. Whether analysis of the full spectrum can recover additional diagnostic information in clinical settings remains largely unexplored.

Lower-leg cellulitis and non-infectious edema provide a demanding test of this clinical utility because they can appear similar while reflecting different physiology and requiring different management. Cellulitis is misdiagnosed in approximately 30% of cases, in part because its signs are nonspecific and no objective test reliably distinguishes it from noninfectious mimics (4–7). Accurate recognition is important: delayed treatment can permit infection to spread to deeper tissues or the bloodstream, whereas overdiagnosis leads to unnecessary treatment and contributes an estimated $195–$515 million in avoidable health-care spending (18, 47). These conditions are also common; skin and soft-tissue infections account for approximately 3% of emergency-department visits, and 70–80% involve the lower extremity (2, 3).

We evaluated whether whole-limb HSI acquired under emergency-department conditions could distinguish cellulitis, non-cellulitis edema, and healthy tissue. We compared classification using standard HSI indices with models that retain the full spectrum, testing whether whole-spectrum analysis recovers diagnostic information lost through spectral reduction. We also performed sensitivity analyses to determine whether classification accuracy varied with Fitzpatrick skin type and whether pigmentation, rather than disease-related physiology, drove model performance as darker skin tones are at higher risk of misdiagnosis and management.

## Methods

### Patient Selection and Cohort Size

We enrolled 98 patients between August 2024 and August 2025 from the emergency department of a regional referral medical center and Level 1 trauma center. All procedures were approved by Lifespan, the Human Research Protection Program of Brown University Department of Emergency Medicine (protocol number 1981418); all participants provided informed consent. Eligible participants were hemodynamically stable adults presenting with lower-extremity swelling who could provide informed consent in English, Spanish, or Portuguese. Patients were excluded for localized rather than uniform edema, inability to image the lower extremity, inability to provide consent, an established diagnosis at screening, pregnancy, limb absence, clinical care needs that prevented study procedures, or prior enrollment. Images were also excluded for low quality, including oversaturation, undersaturation, or large movement artifacts. Healthy participants were recruited to reduce class imbalance. After applying eligibility and image-quality exclusions, 83 patients were included in the final analysis. Demographic and clinical characteristics of the analyzed cohort are summarized in Table 1.

### Imaging System and Acquisition Parameters

Whole-leg hyperspectral images were acquired with an SOC-710-VP hyperspectral camera (Surface Optics Corporation), which records 128 bands from 377 to 1043 nm. Two StarLite® tungsten lamps fitted with StarLite® diffusers provided illumination (19, 20). The lamps flanked the camera and were oriented approximately 45° to the patient’s longitudinal axis to illuminate the legs evenly above the stretcher (Figure 2). Representative images are shown in Figure 3.

**Figure 2.**
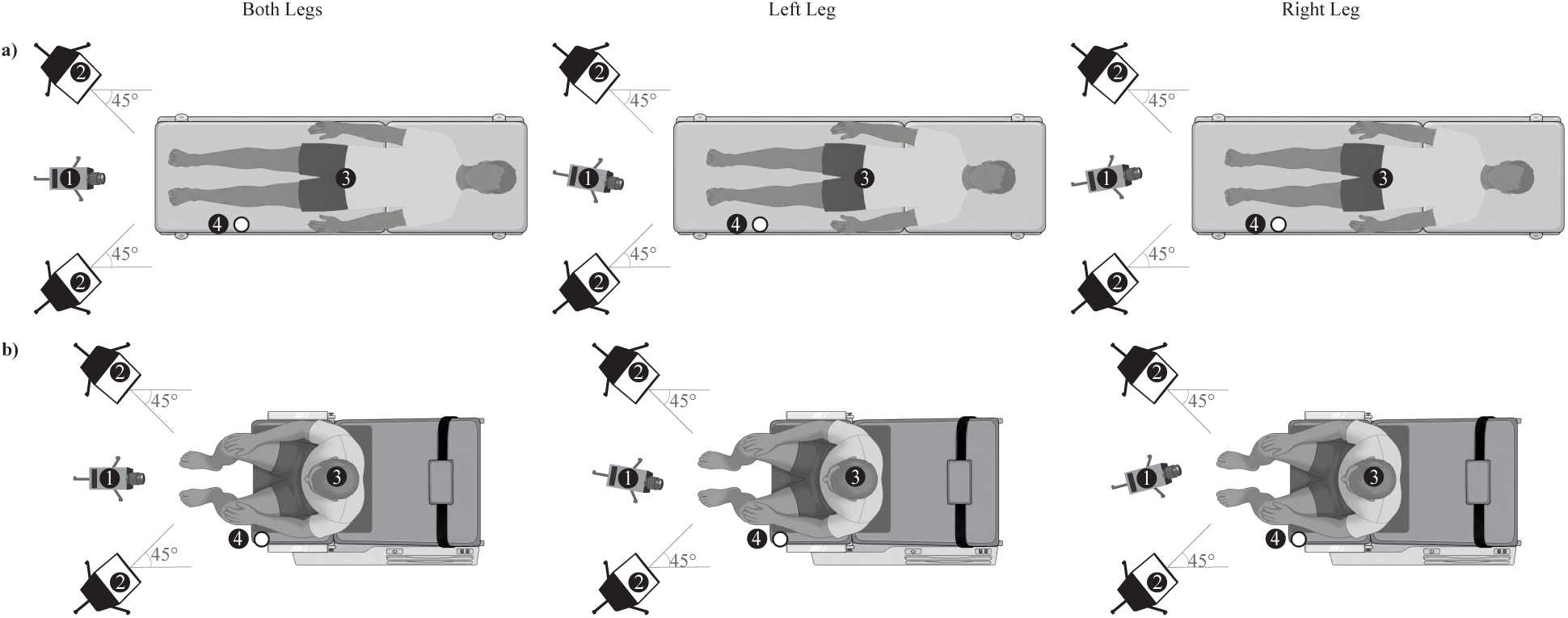
Top-down views of the acquisition setup with the patient (A) supine and (B) seated. Labeled components are: (1) hyperspectral camera, (2) diffused tungsten lamps, (3) patient, and (4) reflectance standard. The camera was rotated on its tripod to acquire three configurations: the left leg, the right leg, and both legs together.

**Figure 3.**
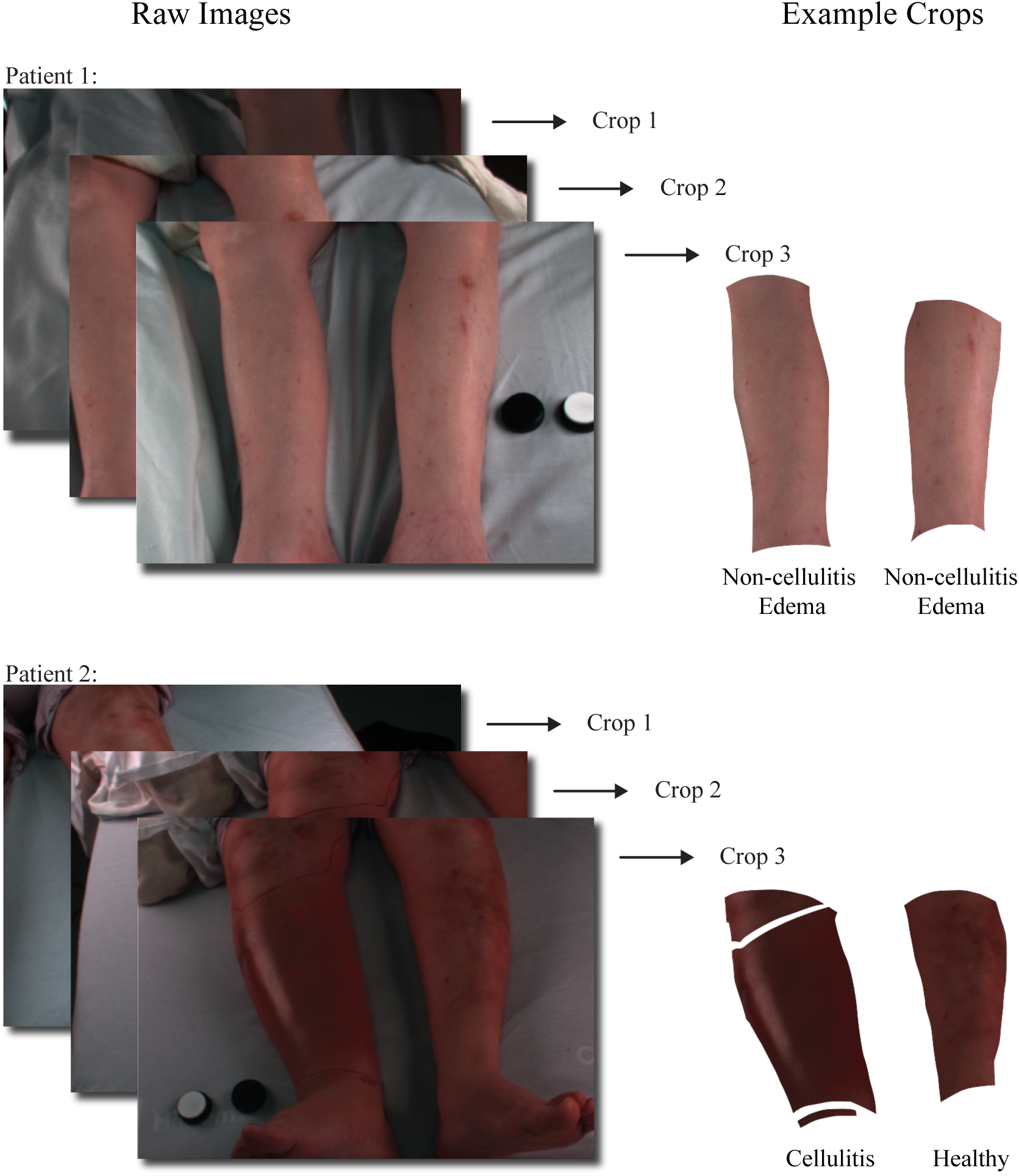
Representative cellulitis and non-cellulitis edema images acquired in the three camera configurations, with an example of the cropped bilateral-leg region. Physician-applied markings delineating disease extent were excluded during cropping; analysis was restricted to lower-leg skin between the knee and foot.

### Image Preprocessing and Labeling

Raw hyperspectral data were calibrated in SRAnalysis software (Surface Optics Corporation) to correct sensor and illumination variability across images. The software first subtracted a dark frame acquired with the optical path blocked, thereby correcting pixel-level offset, sensor noise, and baseline drift. Relative reflectance was then calculated by scaling the dark-corrected radiance at each pixel to a 99% Spectralon reference included in the scene. Equation (1) summarizes this calibration, where the calibrated hypercube is derived from the raw hypercube, dark frame, and Spectralon reference.

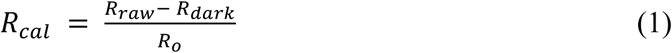

Analyses were restricted to 400–1000 nm, where the silicon detector provided reliable signal-to-noise ratio and calibration (27; Supplementary Figure 2). Residual variation in acquisition geometry, tissue curvature, ambient illumination, and image side could affect spectral intensity after calibration. We therefore applied standard normal variate (SNV) normalization to each spectrum by subtracting its mean and dividing by its standard deviation. SNV recovered previously reported wavelength-dependent sex differences in skin albedo (21), whereas unnormalized spectra produced inconsistent trends (Supplementary Figure 1). All subsequent analyses used SNV-normalized spectra.

Lower-leg regions were manually cropped before labeling and analysis. Crops included visible skin between the knee and ankle while excluding background, clothing, physician-applied markings, and other non-leg regions. Partially imaged legs were retained when the relevant lower-leg tissue was visible. When both legs were captured, each was cropped separately; in unilateral disease, the visible contralateral unaffected leg was retained as a within-patient healthy sample.

Labels were assigned at the leg level from the physician’s diagnosis. All retained tissue pixels from a diagnosed leg were labeled as cellulitis or non-cellulitis edema; pixels from clinically unaffected legs, including eligible contralateral legs from cellulitis and non-cellulitis edema patients, were labeled as healthy controls. The single patient with DVT was not analyzed as a standalone diagnostic class; because disease was unilateral, the contralateral unaffected limb was retained as an additional healthy leg.

### Partial Least Squares (PLS) Based Classification

#### PLS-DA and Variable Importance in Projection (VIP) Scores

We used partial least squares discriminant analysis (PLS-DA), a chemometric classification method well suited to high-dimensional, collinear spectral data (22, 23), to test whether disease-related biochemical changes produced discriminative tissue spectra. PLS-DA projects correlated wavelengths onto a smaller set of latent components while retaining variation associated with class labels. To interpret the fitted models, we calculated variable importance in projection (VIP) scores, which summarize each wavelength’s contribution across components (22). Wavelengths with VIP > 1 were considered influential and were compared with known chromophore absorption features; wavelengths with VIP ≤ 1 were not considered influential.

#### Leave-one-group-out Cross-validation

We evaluated generalization to unseen patients using leave-one-group-out (LOGO) cross-validation, with patient identity as the grouping variable (24–26). In each fold, all observations from one patient were reserved for testing and all remaining patients were used for training. Thus, images and pixels from both legs—and from both healthy and diseased tissue when present—remained in the same fold, preventing leakage of patient-specific spectral features. Data-dependent preprocessing was fit within each training fold and then applied unchanged to the held-out patient.

### Patient-Level Median Analysis

For patient-level analysis, spectra were aggregated by patient and class using the pixel-wise median, yielding one spectrum for each patient–class pair. This reduced pixel noise and local spatial variability. Separate PLS models classified non-cellulitis edema versus healthy tissue, cellulitis versus healthy tissue, and non-cellulitis edema versus cellulitis.

Models were evaluated using patient-grouped nested LOGO cross-validation. Within each outer training set, an inner LOGO loop selected 2–20 PLS components by pooled AUC; ties favored the smaller model. Features were z-score standardized using outer-fold training statistics, and the selected model was then fit to the full outer training set and applied to the held-out patient.

Classification thresholds were selected within each outer training set to maximize the harmonic mean of sensitivity and specificity, then applied to the held-out patient. Predictions were pooled across outer folds to calculate AUC, accuracy, sensitivity, and specificity. Selected component counts and patient-level performance were recorded, and VIP scores were summarized across outer-fold models.

### Pixel-Level Analysis

Pixel-level models treated each preprocessed tissue pixel as an observation, preserving spatial heterogeneity that is lost through patient-level aggregation. Patient-grouped LOGO cross-validation kept all pixels from each patient in the same fold. Separate models classified non-cellulitis edema versus healthy tissue, cellulitis versus healthy tissue, and non-cellulitis edema versus cellulitis using only pixels from the relevant classes.

Pixel-level PLS1 models were fit in R with the bigPLSR package using the SIMPLS algorithm (28, 29). Because nested pixel-level tuning was computationally prohibitive, the number of components was fixed to the mean optimum from the corresponding patient-level median analysis: 6 for non-cellulitis edema versus healthy tissue, 5 for cellulitis versus healthy tissue, and 7 for non-cellulitis edema versus cellulitis. Within each fold, z-score parameters were estimated from the training pixels and applied unchanged to the held-out patient.

Training-fold thresholds maximized the harmonic mean of sensitivity and specificity and were then applied to held-out pixels. Predictions were pooled across folds to calculate AUC, accuracy, sensitivity, specificity, and their harmonic mean. VIP scores were summarized across models, and performance was also stratified by patient and class to assess consistency.

### Spectral Index Calculation and Analysis

Spectral indices were calculated from reflectance spectra using the wavelength bands and derivative-based method of Holmer et al. (30). Reflectance was converted to absorbance by negative logarithmic transformation. THI was the ratio of mean absorbance at 530–590 nm to 785–825 nm; NIR perfusion, 825–925 nm to 655–735 nm; and TWI, 955–980 nm to 880–900 nm. StO₂ was calculated after Gaussian smoothing (6-nm full width at half maximum) and second differentiation as the ratio of minima at 570–590 nm and 740–780 nm. Because the original calibration constants were unavailable, indices were analyzed in raw, unscaled form and represent relative spectral features rather than absolute physiological quantities. This limits comparison with calibrated systems but not within-cohort classification, which depends on relative class differences and uses fold-specific feature scaling.

The four indices were jointly entered into multivariable logistic regression models using the same patient-grouped LOGO framework as the whole-spectrum analyses. Models were fit to both pixel-level values and patient–class medians calculated after index extraction. Within each fold, features were standardized and classification thresholds were selected from the training data by maximizing the harmonic mean of sensitivity and specificity, then applied unchanged to the held-out patient.

### Sensitivity Analysis

We tested whether classification accuracy varied with self-reported Fitzpatrick skin type, recorded as an ordinal predictor from Type 1 to Type 6 (31; Table 2). Because prediction correctness was binary, we modeled correct versus incorrect classification using logistic regression (32). Fitzpatrick type was treated as a linear ordinal predictor, consistent with prior evidence that melanin content correlates with Fitzpatrick skin type (33, 34).

For each classifier, we compared a full model including skin type with an intercept-only reduced model:

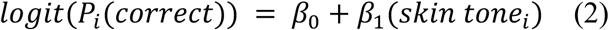

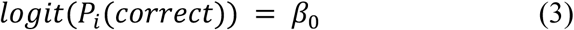

Models were fit by maximum likelihood and compared using a likelihood-ratio test (Equation 4) (35).

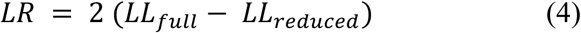

A significant result would indicate that adding skin type improved model fit. The coefficient β₁ represents the change in log-odds of a correct prediction for each one-category increase in Fitzpatrick type, and exp(β_₁_) gives the corresponding odds ratio (36).

Categorical specifications that did not assume a monotonic relationship were explored but not reported because the available events-per-parameter were insufficient for stable estimation (32).

## Results

### Demographics

**Table 1.**
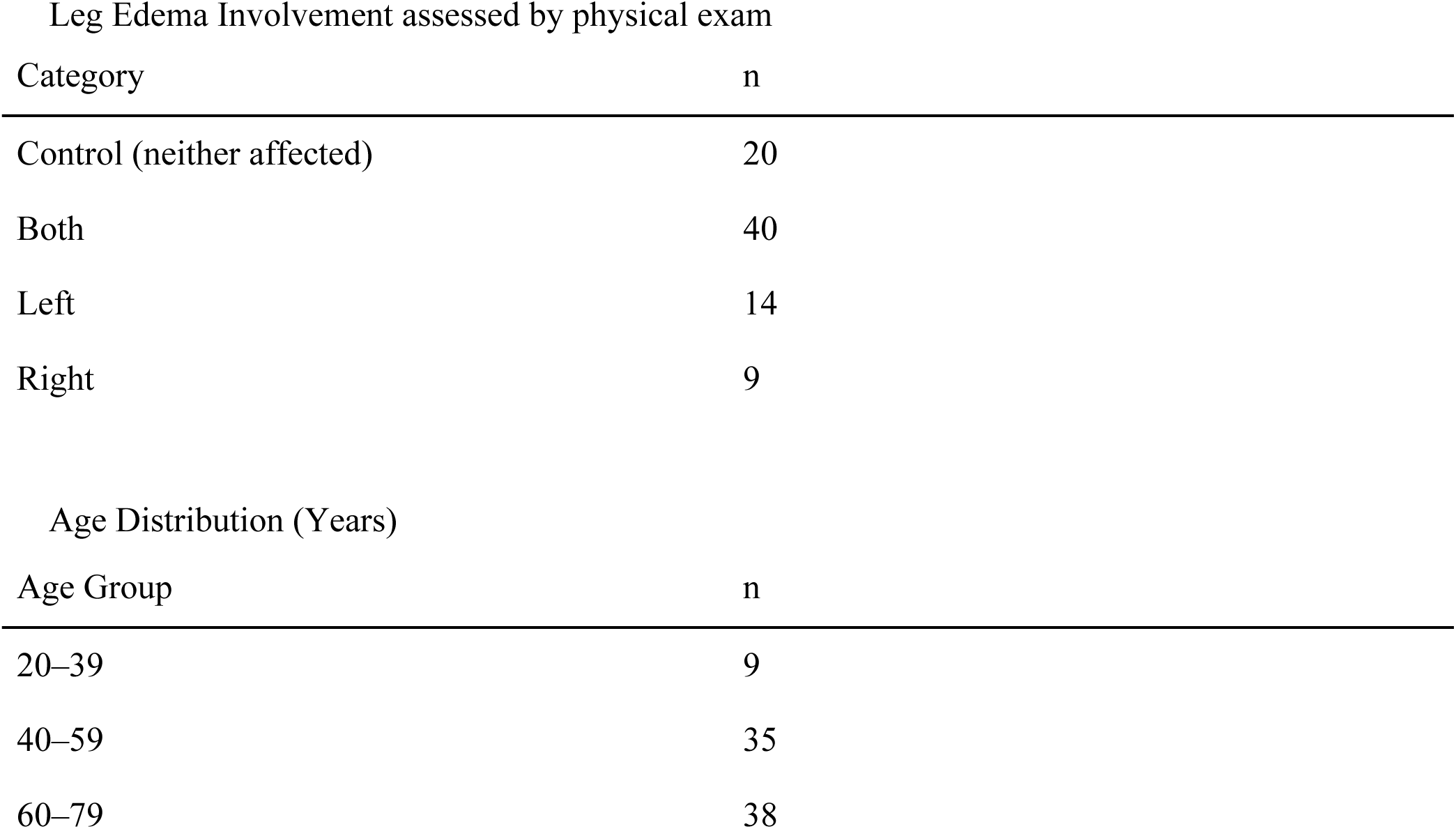
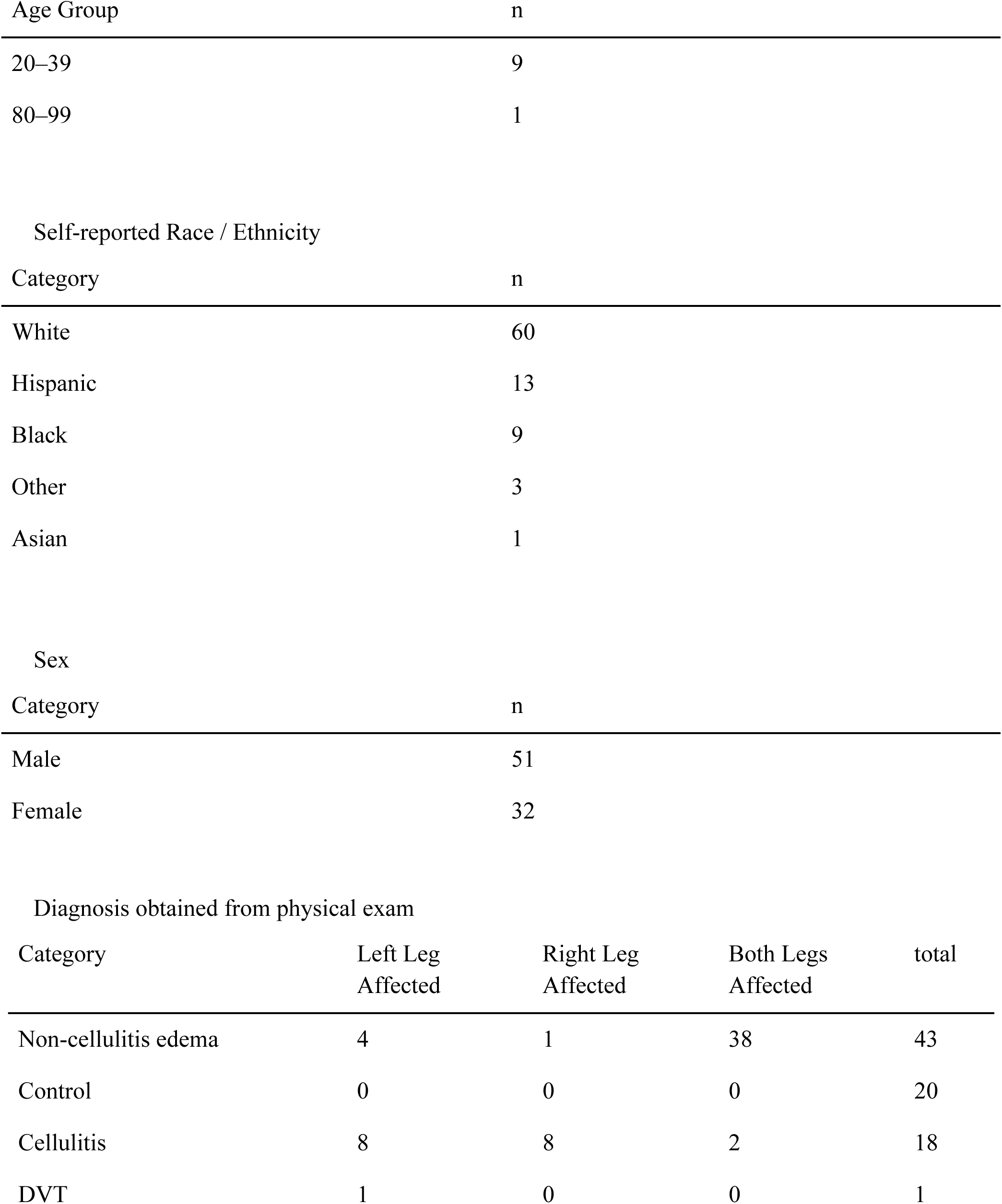
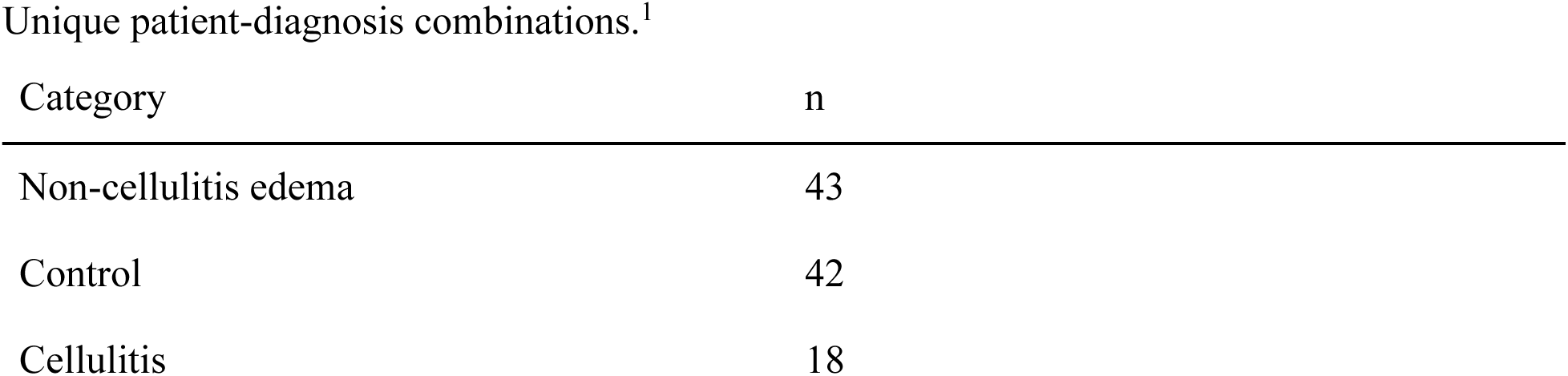
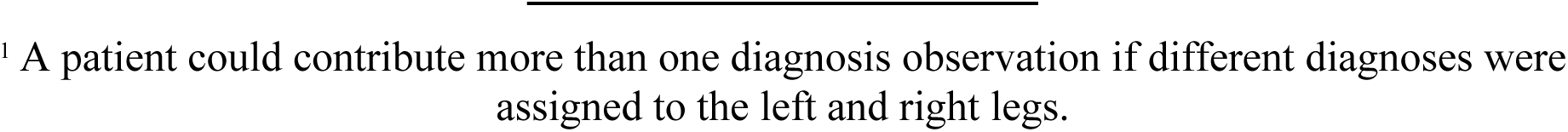
Patient Demographics and Clinical Characteristics (N = 83)

**Table 2.** Self-Reported Fitzpatrick Skin Type.

| Type | Description | n |
| --- | --- | --- |
| 1 | White skin, always burns, never tans | 7 |
| 2 | Fair skin, always burns, tans with difficulty | 22 |
| 3 | Average skin color, sometimes mild burn, tan about average | 28 |
| 4 | Light brown skin, rarely burns, tans easily | 18 |
| 5 | Brown skin, never burns, tans very easily | 8 |
| 6 | Black skin, heavily pigmented, never burns | 0 |
| NA | Prefer not to answer / No response | 0 |

| Category | n |
| --- | --- |
| Enrolled Patients | 98 |
| Excluded Patients | 15 |
| Patients included in analysis | 83 |

### Classification

Figure 4 shows wavelength-wise PLS VIP scores for cellulitis versus healthy, non-cellulitis edema versus healthy, and non-cellulitis edema versus cellulitis. Spatial maps of the whole limb showed spatial variation in NIR perfusion, TWI, StO₂, THI, and PLS scores across the imaged limb (Figure 5). PLS scores varied regionally relative to the classifier threshold, indicating that whole-spectrum models captured heterogeneous disease-associated signal within the analyzed lower-leg region.

**Figure 4:**
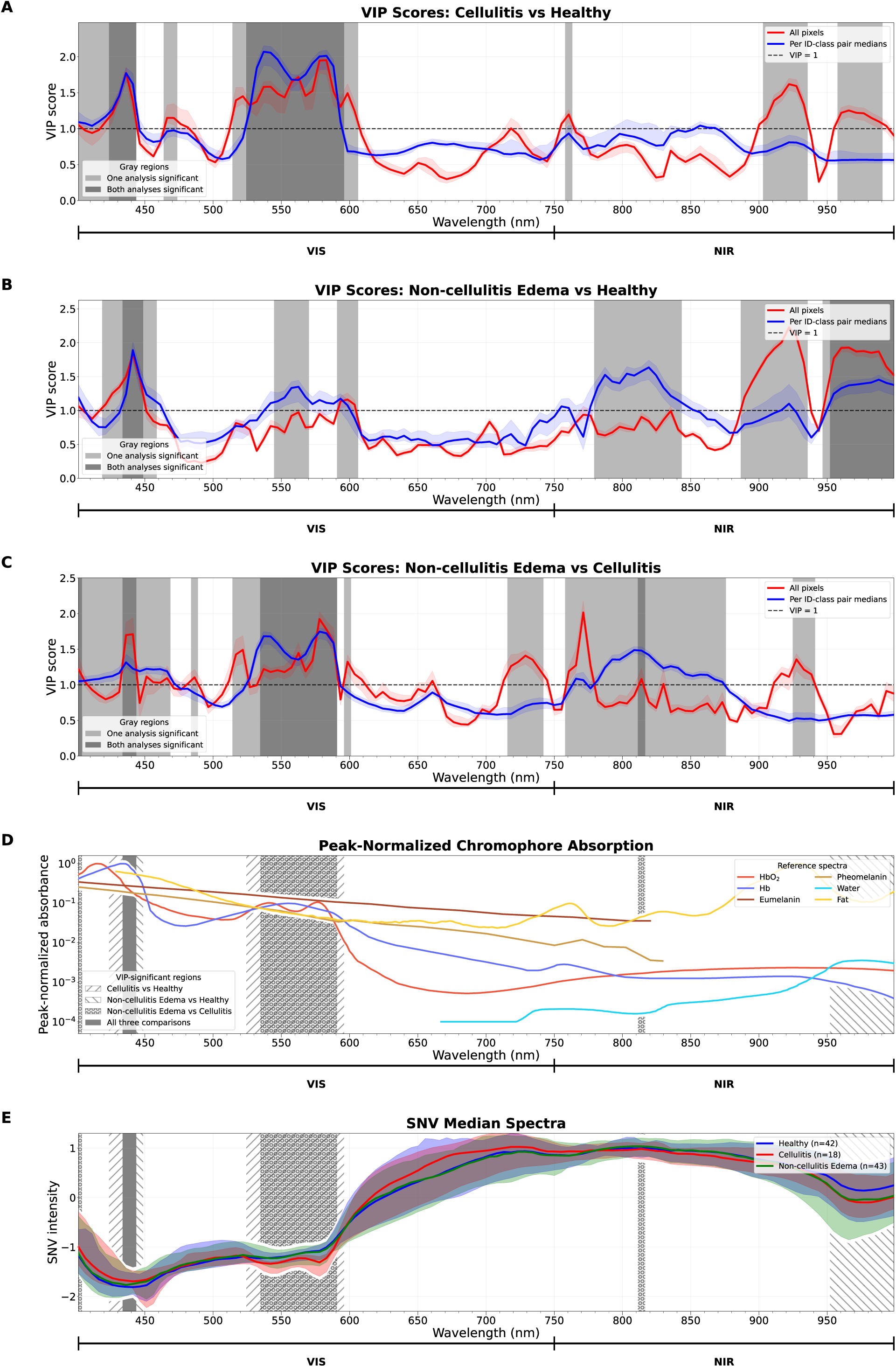
**A–C** show wavelength-wise PLS VIP scores for cellulitis versus healthy, non-cellulitis edema versus healthy, and non-cellulitis edema versus cellulitis, respectively. Red traces represent all-pixel analyses, and blue traces represent patient-level median analyses. Solid lines show median VIP scores across outer folds, shaded bands indicate the 2.5th–97.5th percentile range, and the dashed line marks VIP = 1. Gray regions indicate influential wavelengths (VIP > 1 across the full percentile interval), with darker shading denoting agreement between both analyses. **D** shows peak-normalized chromophore and absorber spectra for comparison with VIP-significant regions. Hatched regions mark wavelengths significant in both all-pixel and patient-level analyses for a given task, while solid dark gray regions indicate wavelengths significant across all three classification tasks. **E** shows SNV-normalized median spectra for healthy, cellulitis, and non-cellulitis edema groups. Shaded bands indicate the 2.5th–97.5th percentile range across patient-level median spectra, and the VIP-significant regions from panel D are overlaid for reference.

**Figure 5:**
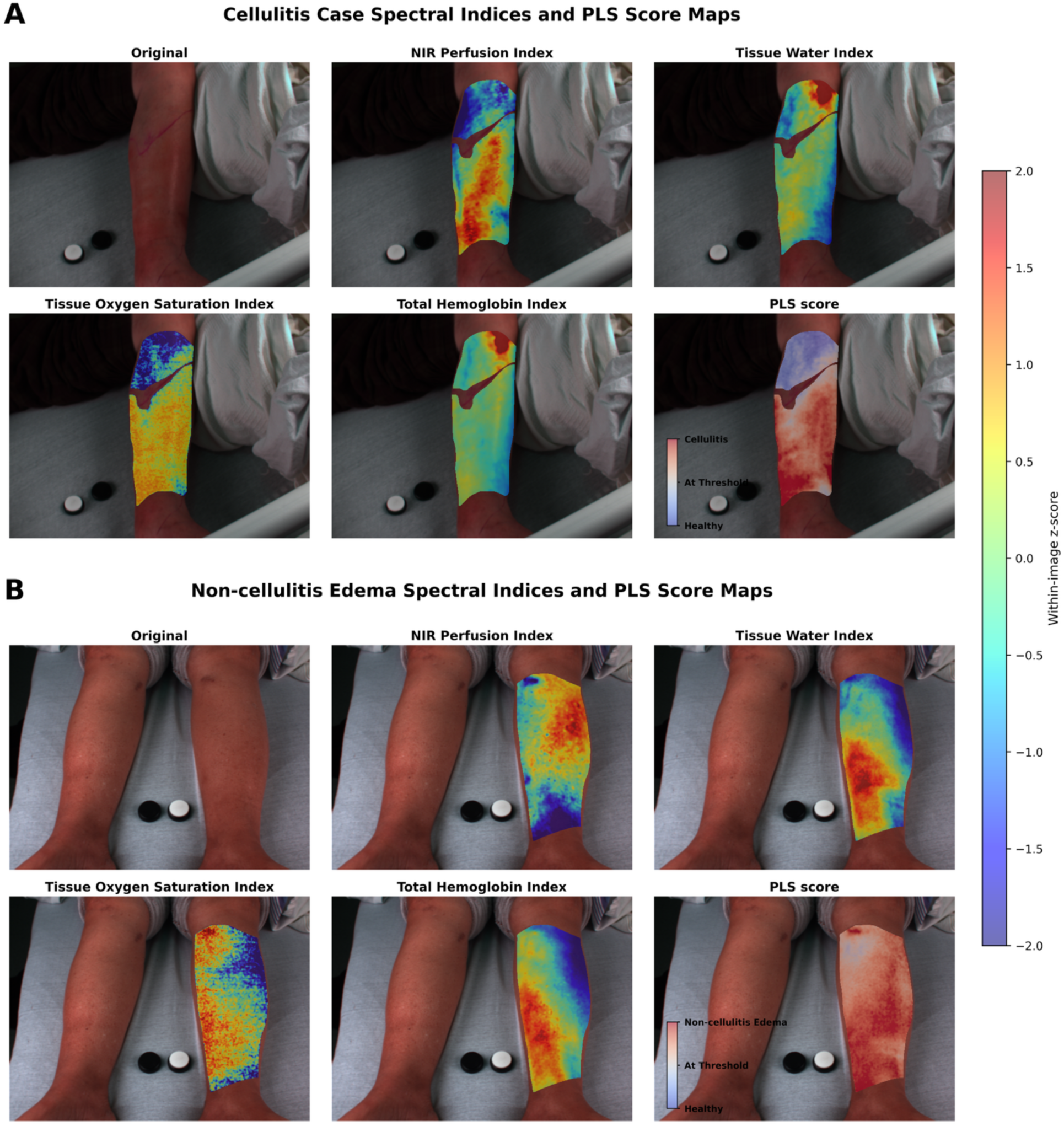
Spectral Indices for Cellulitis and Non-cellulitis Edema. Representative RGB images from patients with (A) cellulitis and (B) non-cellulitis edema are shown with NIR perfusion, tissue water, tissue oxygen saturation, total hemoglobin, and PLS score overlays. The four index overlays represent relative perfusion, water content, hemoglobin oxygenation, and hemoglobin abundance, respectively, and are displayed as within-image z-scores. PLS overlays show raw scores shifted relative to each classifier’s decision threshold, so values below, at, and above zero correspond to the healthy side, threshold, and disease side of the classifier.

Across tasks, classification was strongest for comparisons involving cellulitis (Figure 6). At the patient level, cellulitis versus healthy achieved high AUCs with both derived indices and whole-spectrum features (0.96 and 0.93, respectively). Non-cellulitis edema versus cellulitis was similarly strong for both approaches (0.89 and 0.92). McNemar’s exact test showed no significant difference in paired classification correctness between index-based and whole-spectrum models for either cellulitis versus healthy (patient-level p = 1.0; pixel-level p = 1.0) or non-cellulitis edema versus cellulitis (patient-level p = 0.8; pixel-level p = 0.6), suggesting that standard indices captured much of the cellulitis-related signal.

**Figure 6:**
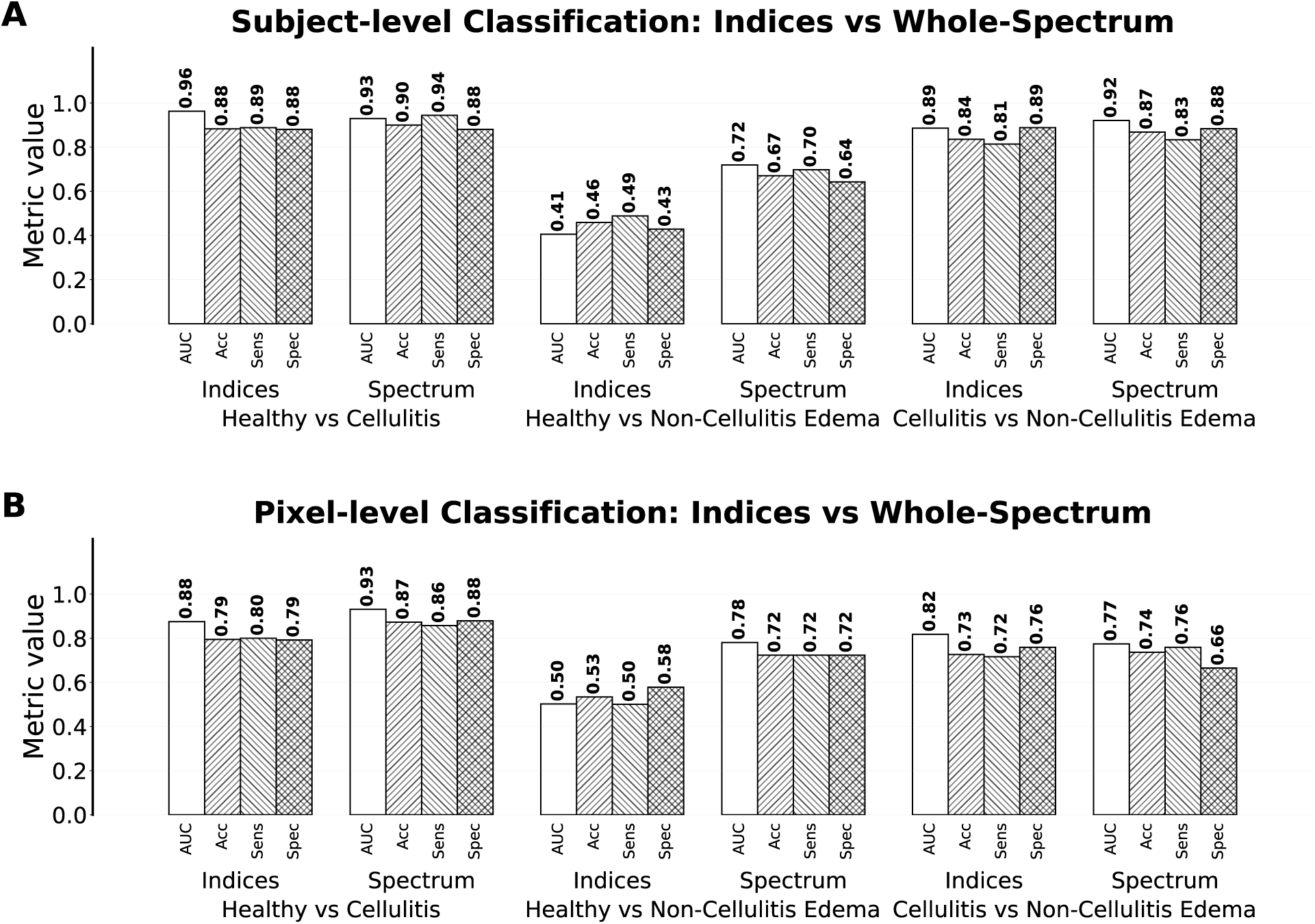
Bar plots compare pooled classification performance across three binary tasks: cellulitis vs healthy, non-cellulitis edema vs healthy, and non-cellulitis edema vs cellulitis. **A** shows patient-level performance after measurements were summarized as patient-class medians, with one median feature representation per patient and class. **B** shows pixel-level. Within each task, models using derived HSI indices are compared with whole-spectrum PLS models. Bars show AUC, accuracy, sensitivity, and specificity, with metric values labeled above each bar. For the non-cellulitis edema vs cellulitis comparison, non-cellulitis edema was treated as the positive class.

A supplementary unilateral-cellulitis analysis tested whether models assigned the same prediction to both limbs of the same patient. Same-class prediction rates were not higher than expected from each model’s overall sensitivity and specificity, supporting the interpretation that cellulitis classification was not driven primarily by patient-level or acquisition-level factors shared across limbs.

Non-cellulitis edema versus healthy classification showed the clearest benefit of whole-spectrum modeling (Figure 6). At the patient level, the index-based model performed poorly (AUC 0.41), whereas the whole-spectrum model improved performance (AUC 0.72); paired correctness was significantly higher for the whole-spectrum model (McNemar p = 0.003). The same pattern appeared at the pixel level, where AUC improved from 0.50 with indices to 0.78 with whole-spectrum features, again with significantly higher paired correctness (p = 0.03). Thus, unlike cellulitis-related comparisons, edema-versus-healthy discrimination depended more strongly on information distributed across the full spectrum than on predefined HSI indices.

Pixel-level models generally performed similarly to or below patient-level median models for cellulitis-related tasks (Figure 6). For cellulitis versus healthy, whole-spectrum AUC remained high at the pixel level (0.93), but index performance decreased from 0.96 to 0.88. For non-cellulitis edema versus cellulitis, whole-spectrum AUC decreased from 0.92 at the patient level to 0.77 at the pixel level. These results suggest that patient-level summaries provide a more stable representation of disease-associated spectral differences than individual pixels. For comparison, Figure 7 shows the discrimination performance of selected spectral indicies computed using logistic regression.

**Figure 7:**
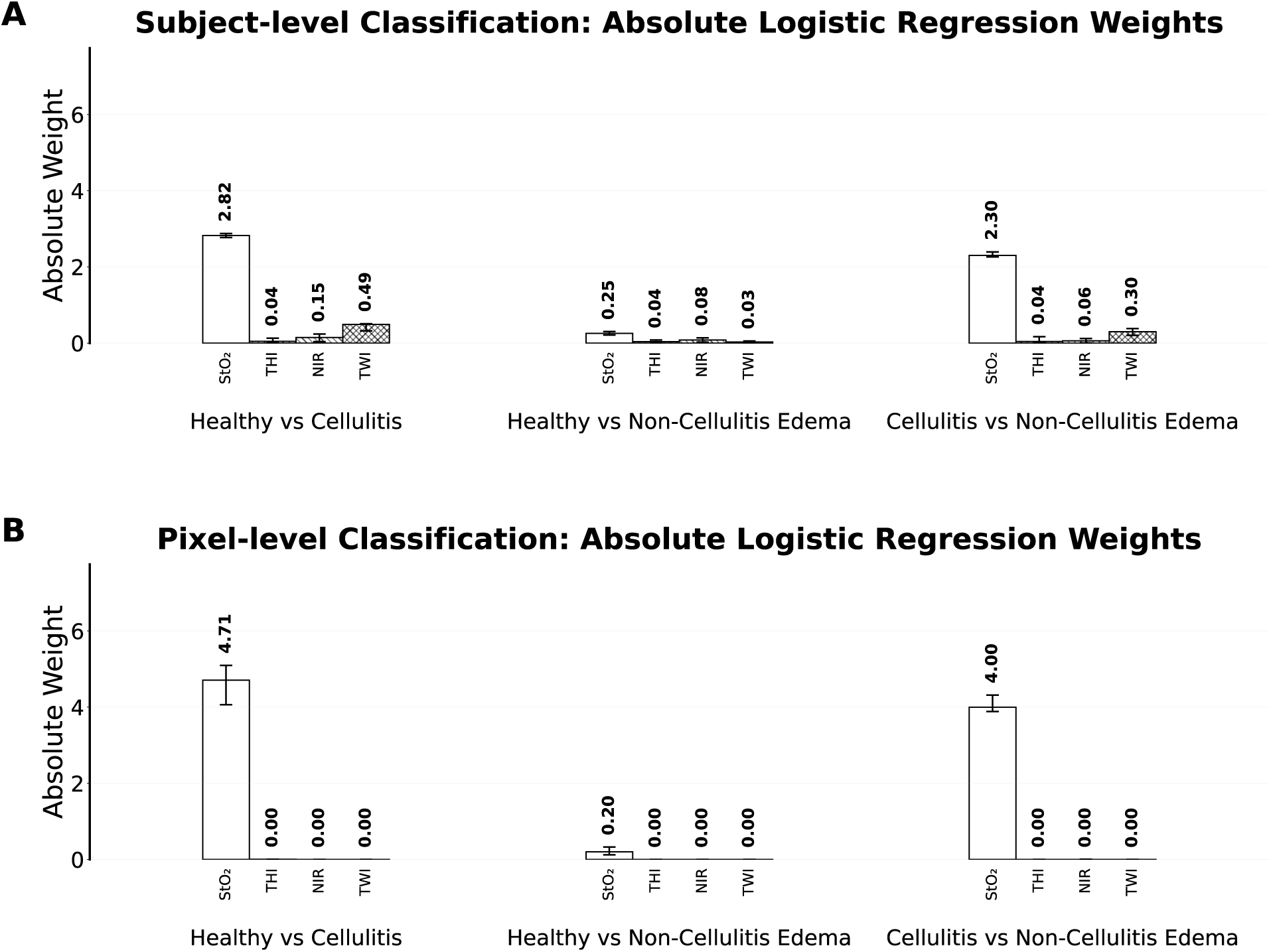
Indices logistic regression results. **A,** Patient-level models using median feature values per ID/class pair. **B,** Pixel-level models. Bars show the mean absolute standardized logistic regression coefficient for each HSI-derived index across LOGO folds. Each classifier was evaluated using four derived indices: oxygen saturation index (StO₂), tissue hemoglobin index (THI), near-infrared perfusion index (NIR), and tissue water index (TWI). Error bars indicate the 2.5th–97.5th percentile range of absolute coefficient values across LOGO folds. Larger absolute coefficients indicate greater relative contribution to the classifier after feature standardization, independent of coefficient direction.

### Sensitivity Analysis

Sensitivity analyses found no significant association between Fitzpatrick skin type and classification accuracy for any model (Table 3). Effect estimates were small and generally close to null; most coefficients were positive, with one exception for patient-level cellulitis versus healthy classification. These results suggest that SNV-normalized spectral features were not primarily driven by pigmentation across the Fitzpatrick types represented in this cohort. However, no participants reported Fitzpatrick Type 6, so performance in more heavily pigmented skin could not be assessed.

#### Sensitivity Analysis

**Table 3.**
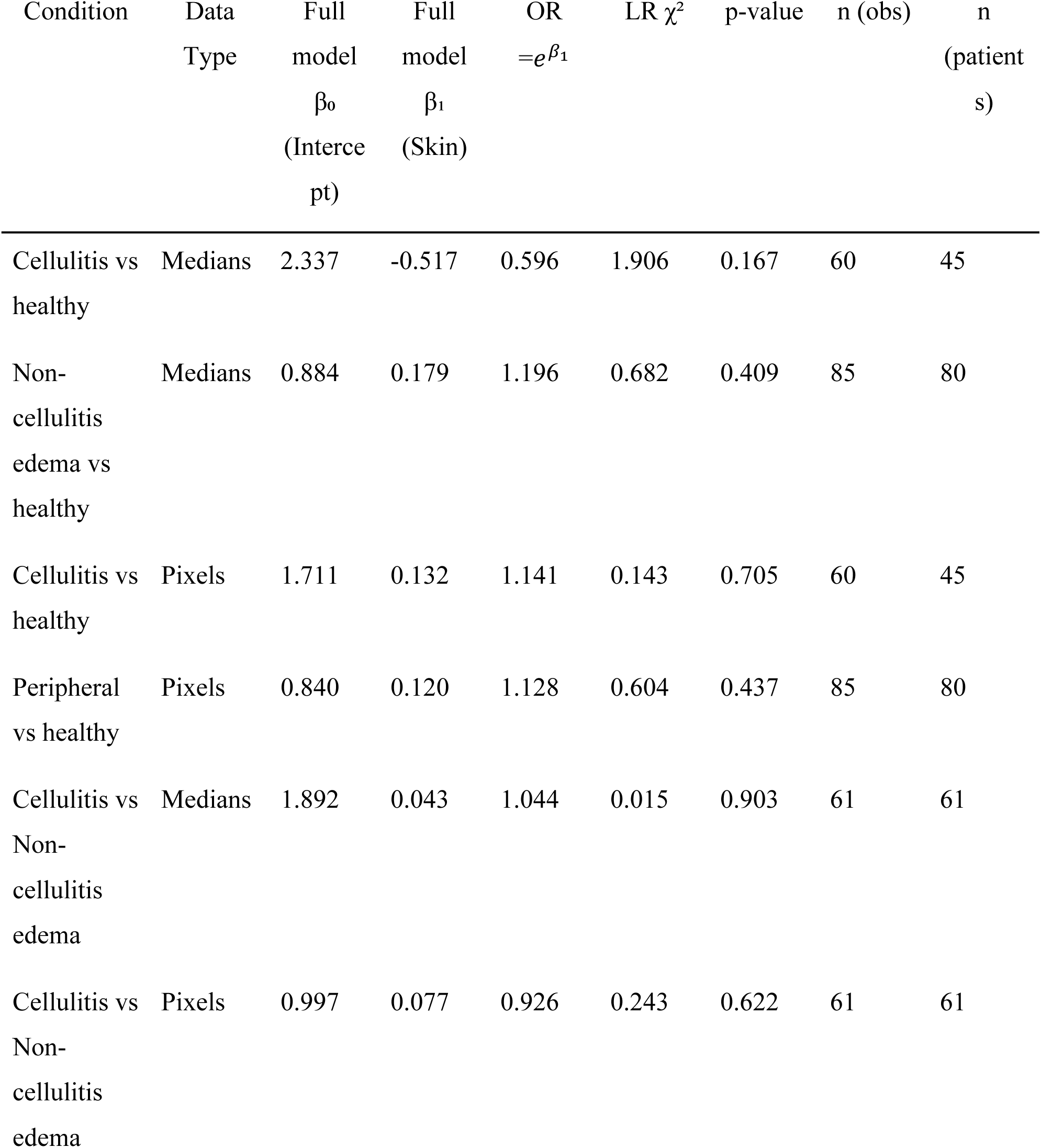
Sensitivity Analysis results.

Most models showed a positive association between higher Fitzpatrick skin type and classification accuracy, with one exception; however, all effect sizes were small.

## Discussion

In this paper, we evaluated hyperspectral imaging (HSI) of whole limbs as a diagnostic tool in the emergency department of lower-leg cellulitis and edema. We compared the full-spectrum analysis provided by HSI against standard spectrum indices. Standard indices were sufficient to capture the cellulitis signal, whereas full-spectrum modeling improved edema-versus-healthy discrimination. Skin tone did not affect diagnostic accuracy in this dataset. These results support further study of HSI as a point-of-care diagnostic tool and suggest that retaining full-spectrum information may complement interpretable clinical indices.

### Classification

Visible/near-infrared HSI captured physiologically meaningful differences among cellulitis, non-cellulitis edema, and healthy lower-extremity tissue. Classification was strongest for comparisons involving cellulitis, where derived HSI indices and whole-spectrum models performed similarly. This pattern is consistent with cellulitis as an inflammatory process characterized by erythema, hyperemia, and altered oxygenation (39, 40). StO₂ was the dominant index in cellulitis-related classifiers, and VIP scores highlighted visible hemoglobin absorption regions near approximately 450 nm and 530–600 nm, consistent with oxyhemoglobin and deoxyhemoglobin features (41). Together, the classification, logistic-weight, and VIP results suggest that oxygenation-and perfusion-related indices capture much of the discriminatory signal associated with cellulitis, while the unilateral cellulitis analysis argues against model performance being driven primarily by shared patient-level or acquisition-level factors.

Unlike cellulitis, non-cellulitis edema is not primarily an inflammatory hyperemia process; it reflects excess interstitial fluid and associated changes in tissue optical properties. Derived indices performed poorly for non-cellulitis edema versus healthy classification, with small logistic regression weights across all four indices, while whole-spectrum models substantially improved performance (Figure 7). Median spectra showed differences near water-and fat-associated features, consistent with prior diffuse reflectance work on dermal water content (42–44). However, these features are broad and overlapping in the visible/NIR range, unlike the stronger water, lipid, and collagen absorption features available in SWIR imaging (45); whole-spectrum analysis may therefore have shown improved discrimination ability by integrating multiple weak spectral-shape and scattering signals rather than relying on a single predefined index such as TWI.

The non-cellulitis edema versus cellulitis comparison also showed wavelength importance near 830 nm. Although this region may include cytochrome c oxidase-related absorption, overlap from hemoglobin, scattering, tissue composition, vascular structure, and acquisition geometry prevents attribution to a single chromophore (46). More broadly, this finding suggests that whole-spectrum models capture information beyond standard oxygenation and perfusion indices.

Patient-level median models generally provided more stable or stronger performance than pixel-level models. Aggregation may reduce local variability from heterogeneous disease presentation, hair follicles, vasculature, pigmentation, motion, and image blur (47, 48). Physician-selected regions also likely contained mixtures of affected and relatively normal tissue while assigning every pixel the same diagnostic label. Patient-class medians may therefore better represent each patient’s overall spectral phenotype. Future pixel-level work with more precise annotations should account for unequal region sizes and correlations among adjacent pixels.

Finally, although the sensitivity analysis did not identify a significant association between Fitzpatrick skin type and classification performance within the represented range, larger and more diverse cohorts are needed to determine whether these spectral patterns generalize across pigmentation, disease severity, chronicity, imaging conditions, and clinical presentation. HSI could provide important diagnostic information for equitable and high-quality care across different skin types to avoid misdiagnosis and incorrect treatment.

## Conclusion

Visible/NIR HSI acquired in the emergency department distinguished cellulitis, non-cellulitis edema, and healthy lower-leg tissue despite real-world acquisition variability. Standard HSI indices captured much of the cellulitis-related signal, but whole-spectrum analysis improved discrimination of non-cellulitis edema from healthy tissue, indicating that clinically useful information can be lost through spectral reduction. Classification accuracy was not significantly associated with Fitzpatrick skin type within the range represented in this cohort. These findings support further evaluation of HSI as a rapid, noninvasive point-of-care tool and of full-spectrum analysis as a complement to standard clinical HSI indices.

## Data Availability

All data produced in the present study are available upon reasonable request to the authors

## Acknowledgements

We thank the patients and the staff of the Brown University Emergency Department, and the Brown University Emergency Department for providing funds supporting this work. We thank Nora Sachse for pilot work setting up the camera. This research was supported in part by the Intramural Research Program of the National Institutes of Health (NIH, 1ZIAEY000558 to BRC). The contributions of the NIH authors are considered Works of the United States Government. The findings and conclusions presented in this paper are those of the authors and do not necessarily reflect the views of the NIH or the U.S. Department of Health and Human Services.

## Code/Data Availability

Data and code will be made available upon reasonable request.

