## Supplementary Information for "Hyperspectral imaging in the emergency department to characterize lower leg edema"

**Supplementary Materials**

**Unilateral sensitivity analysis**

To assess whether models captured unilateral disease rather than assigning the same prediction to both sides of the same patient, we further analyzed 15 unilateral cellulitis IDs with both cellulitis and healthy samples; this analysis focused on cellulitis because most non-cellulitis edema cases were bilateral. Same-class prediction was defined as exactly one side being correct, which in a binary unilateral case indicates that both sides were assigned the same class. Same-class rates were compared with expected rates derived from each model’s overall sensitivity and specificity; this expected rate assumes that the model performs about as well on unilateral cases as it does overall, and that getting one side right does not directly change the chance of getting the other side right. Same-class prediction rates were 33.3% for all-pixel indices, 40.0% for all-pixel whole-spectrum features, 33.3% for patient-level median indices, and 33.3% for patient-level median whole-spectrum features. These rates were not significantly different from sensitivity/specificity-based expectations for all-pixel indices (expected 32.6%, p = 1.0), all-pixel whole-spectrum features (expected 22.6%, p = 0.1), patient-level median indices (expected 20.4%, p = 0.2), or patient-level median whole-spectrum features (expected 16.6%, p = 0.09). Logistic regression weights from the index-based models further indicated that StO₂ contributed most strongly to cellulitis-related classification, with the largest absolute weights observed for cellulitis versus healthy and non-cellulitis edema versus cellulitis at both the patient and pixel levels.

**Healthy Patient Albedo Stratified by Sex**

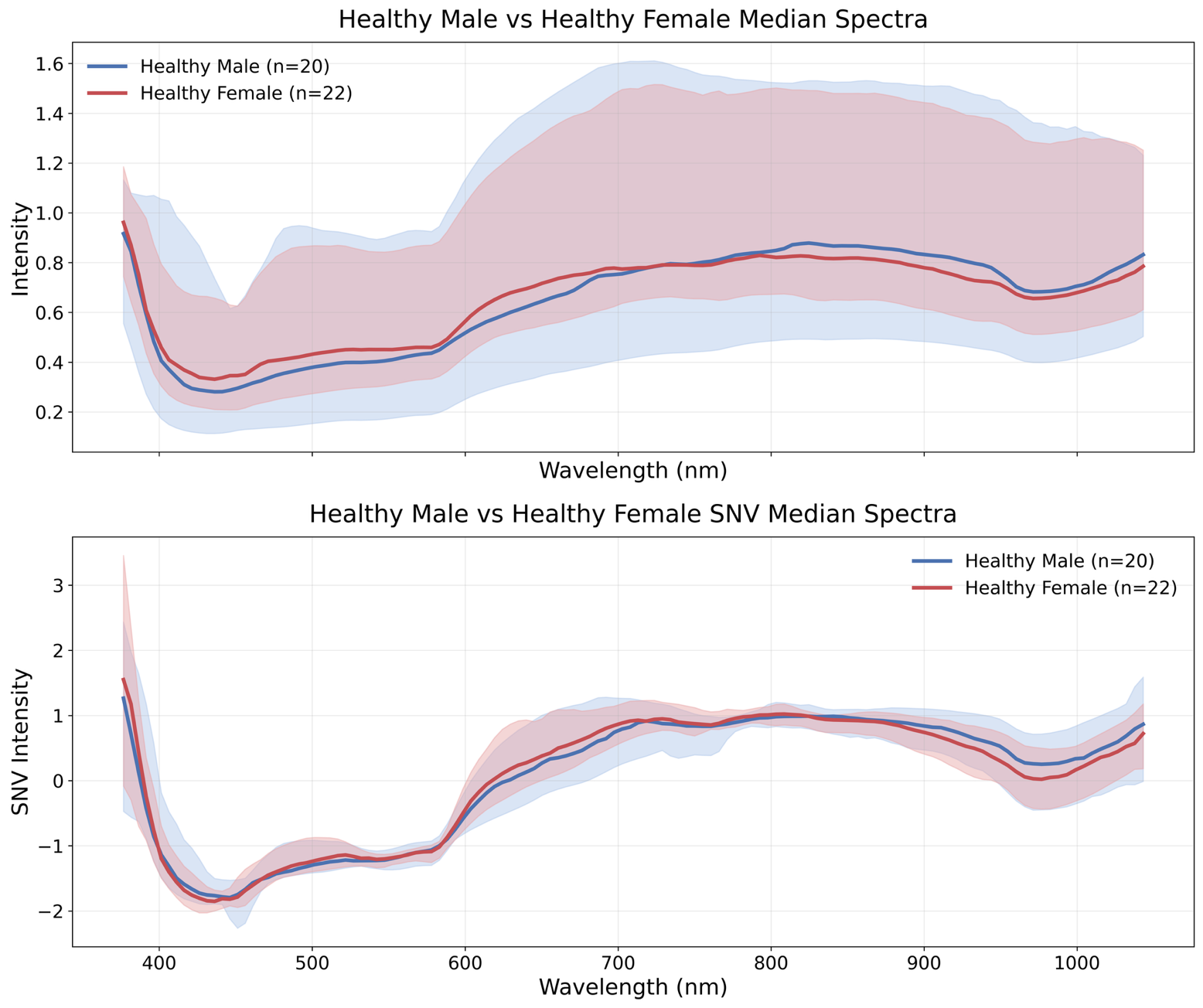

**Supplementary Figure 1**: Sex-stratified median albedo spectra from healthy patients after white-point calibration. Top: Albedo spectra prior to SNV normalization. Bottom: Albedo spectra after SNV normalization. Shaded regions represent the 2.5th–97.5th percentile range across patient-level median albedos. After SNV normalization, males show relatively higher albedo from approximately 400–500 nm, followed by a reversal near 1000 nm, a pattern consistent with Tsai et al. (2021) (21). This trend is not present in the unnormalized spectra.

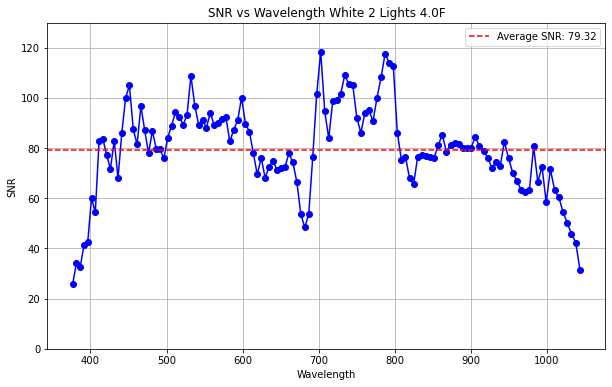

**Supplementary Figure 2**: SNR vs Wavelength chart shows that SNR decreases greater than 1000 and less than 400.
